# Estimating the Final Epidemic Size for COVID-19 Outbreak using Improved Epidemiological Models

**DOI:** 10.1101/2020.04.12.20061002

**Authors:** Rajesh Ranjan

## Abstract

Final epidemic sizes of different geographical regions due to COVID-19 are estimated using logistic, SIR and generalized SEIR models. These models use different parameters which are estimated using non-linear fits from the available data. It is found that both SIR and generalized SEIR models give similar estimations for regions where curves show signs of flattening. A study of these models with data from China indicates that in such cases these estimates may be more reliable. However, recent trend indicate that unlike China, the decline in infection rate for the US and other European countries is very slow, and does not follow a symmetric normal distribution. Hence a correction is introduced to account for this very slow decline based on the data from Italy. The estimates with all these models are presented for the most affected countries due to COVID-19. According to these models, the final epidemic size in the US, Italy, Spain, and Germany could be 1.1, 0.22, 0.24 and 0.19 million respectively. Also, it is expected that curves for most of the geographical regions will flatten by the middle of May 2020.

## 1 Introduction

COVID-19 is caused by novel coronavirus SARS-CoV-2, for which there is no specific medication or vaccine approved by medical authorities. This disease is transmitted by inhalation or contact with infected droplets or fomites, and the incubation period may range from 2 to 14 days[1]. Though, the case fatality rate is estimated to range from 2 to 3%, the disease can be fatal to older adults (about 27% for 60+ age groups) and those with an underlying co-morbid conditions[2].

In the recent days, Coronavirus disease 2019 (COVID-19) has emerged as an unprecedented challenge before the world. As of April 8, 2020, there have been about 1.5 million confirmed cases of COVID-2019 and about 90,000 reported deaths globally. For subsequent discussions, the default year is 2020 for all the dates mentioned unless otherwise specified.

The global increase in the number of cases every day can be seen in Fig. 1(a). Compared to around 0.8 million cases on March 30, the case has been doubled in 10 days. The nature of growth can be better seen on a logarithmic scale as shown in Fig. 1(b). A closer examination shows three periods of exponential growth with different exponents. In order to examine these periods, the growth in some of the most affected countries is shown in Fig. 2. Examining Fig. 1(a) along with Fig. 2 provides interesting insights.

**Figure 1:**
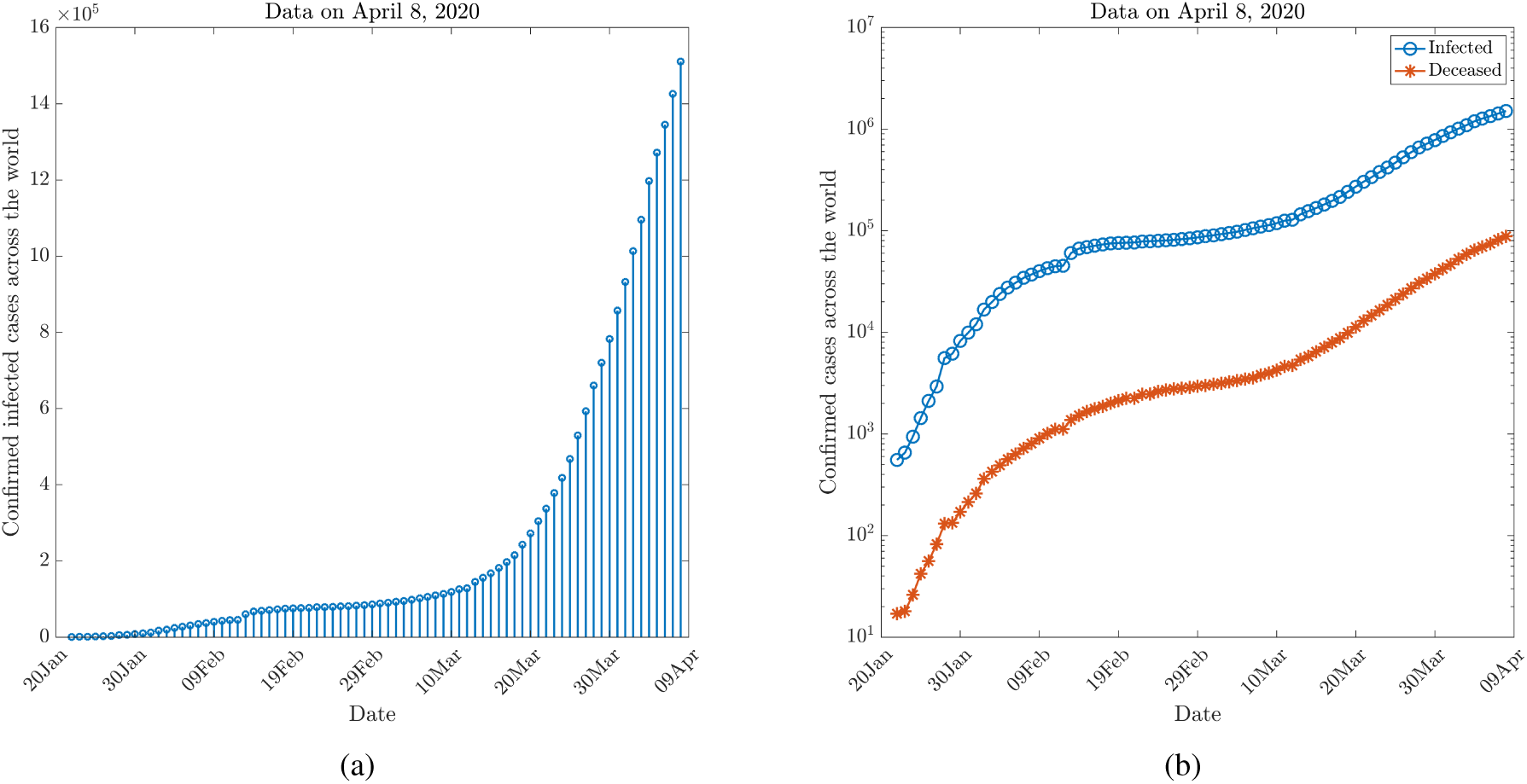
Confirmed COVID-19 cases across the world.

**Figure 2:**
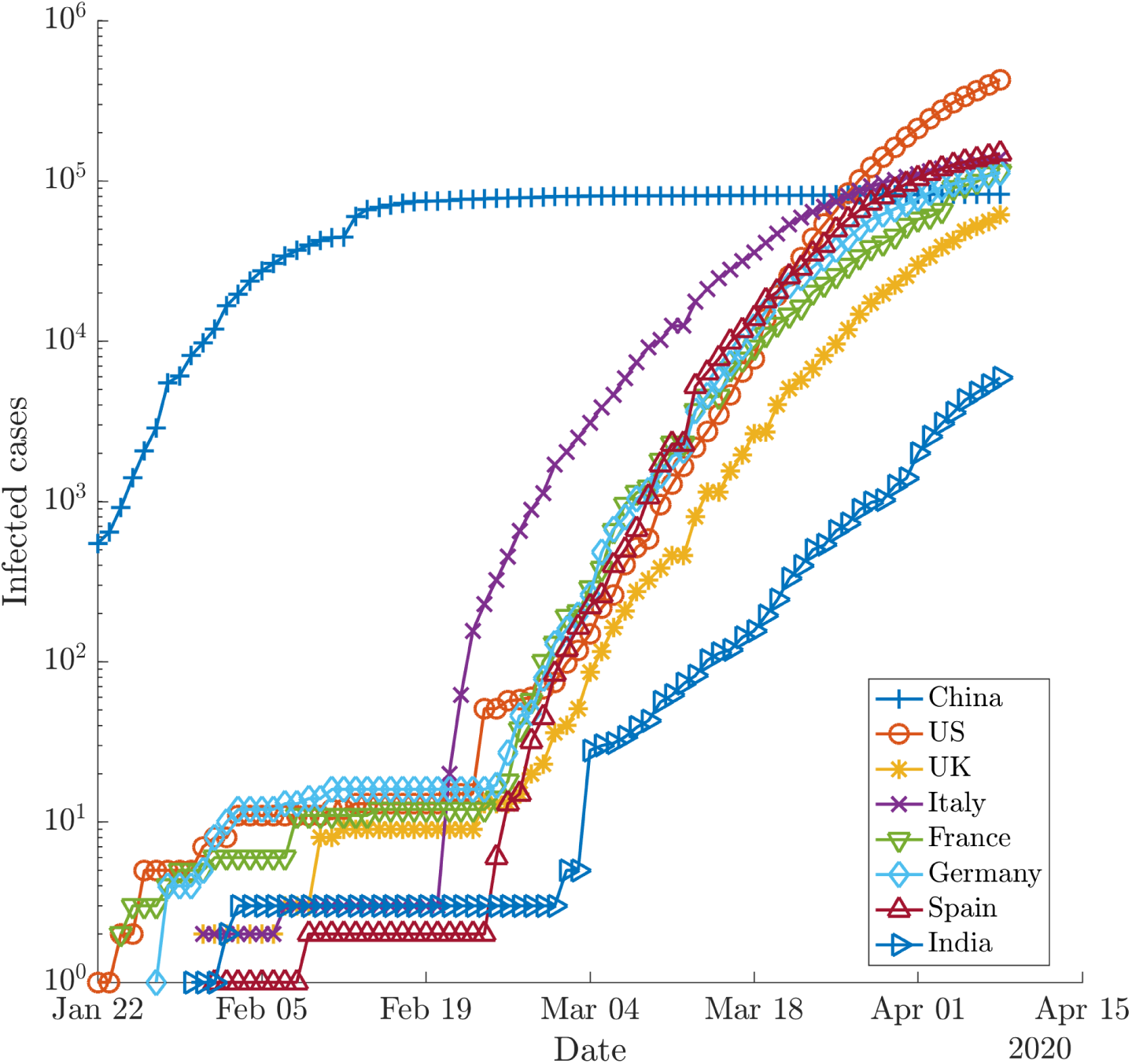
Confirmed COVID-19 cases in most affected countries and India.

The first rapid growth corresponds to cases between Jan 22 to the end of January and indicates the growing cases in China. The plateau from the end of January to Feb 10 indicates the effects of strict lockdown imposed in China to control the spread. During this period, the rest of the world had a very few officially confirmed cases. From mid of February till March 10, a slow exponential growth corresponds to COVID-19 cases spreading slowly in the rest of the world. During this period, the countries most affected by the epidemic are South Korea, Iran, Italy, etc. A third rapid exponential growth begins around March 11, when the epidemic spreads to the United States and other European countries such as Spain, France, Germany and the UK. Countries like India exhibit a very slow growth during the entire period, which may correspond to early suspension of international flights and strict social distancing measures.

Fig. 1(a) shows the number of worldwide deaths due to this epidemic, which also shows very similar distribution as the infected cases as expected. The death percentage is about 6% of the total infections. However, this proportion is different for different geographical regions and depends on the demography of population. In order to further characterize the spread of COVID-19 in different regions, we define two ratios:

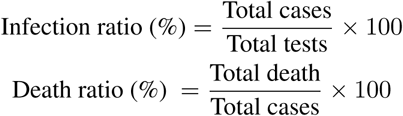

These ratios are presented in Table 1 based on data till April 8. The infection ratio (IR) is highest in Spain among these countries (45%) and least in India (4%). US has about 20% IR, but there is a high disparity in this value for different states. For example, New Jersey and New York have IR > 40%, whereas for Washington it is just about 10%. Death ratio (DR) is also different in these regions listed. Italy reports highest DR of about 13%, while the lowest death ratio is reported from Germany (∼ 2%).

**Table 1:**
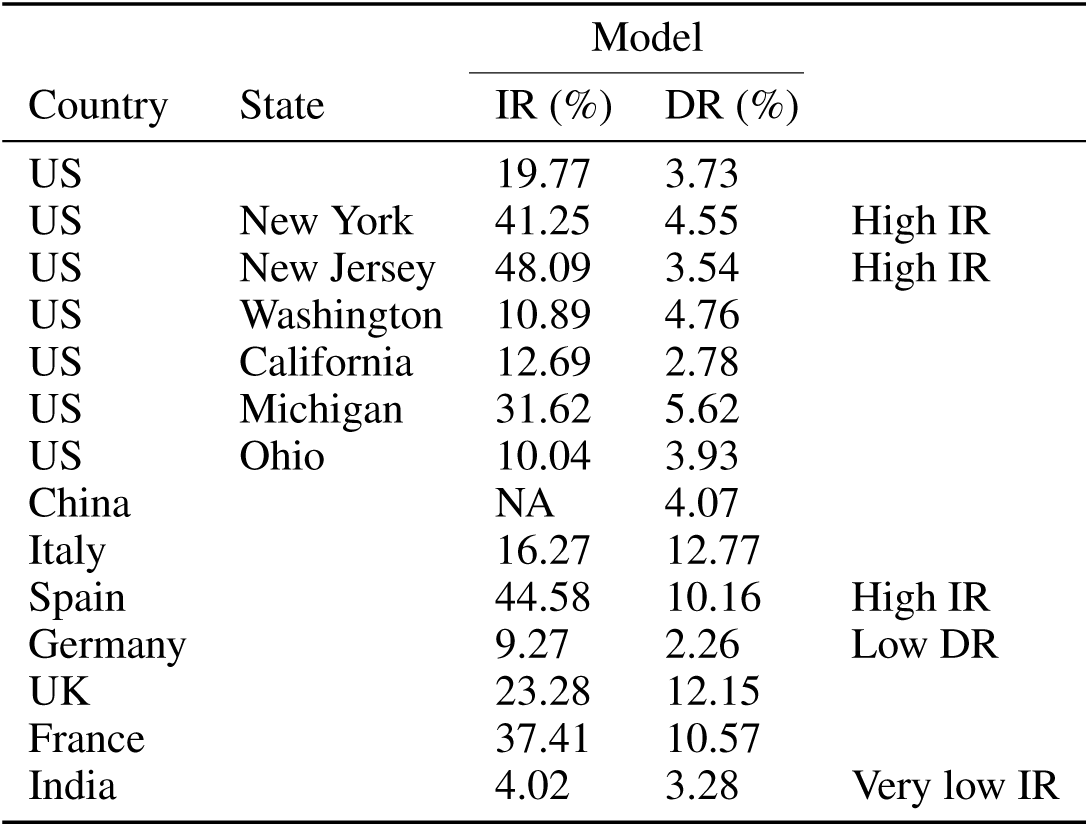
Characteristics of spread in different countries

| Country | State | Model |  |  |
| --- | --- | --- | --- | --- |
|  |  | IR (%) | DR (%) |  |
| US |  | 19.77 | 3.73 |  |
| US | New York | 41.25 | 4.55 | High IR |
| US | New Jersey | 48.09 | 3.54 | High IR |
| US | Washington | 10.89 | 4.76 |  |
| US | California | 12.69 | 2.78 |  |
| US | Michigan | 31.62 | 5.62 |  |
| US | Ohio | 10.04 | 3.93 |  |
| China |  | NA | 4.07 |  |
| Italy |  | 16.27 | 12.77 |  |
| Spain |  | 44.58 | 10.16 | High IR |
| Germany |  | 9.27 | 2.26 | Low DR |
| UK |  | 23.28 | 12.15 |  |
| France |  | 37.41 | 10.57 |  |
| India |  | 4.02 | 3.28 | Very low IR |

In order to have further insights into the spread of this epidemic, it is important to include the effects of social distancing. The basic reproduction number *R*_0_ for SARS-CoV-2 can be as high as 4, indicating a very high transmission rate of the disease. Therefore, in the absence of any available treatment approved by medical authorities and World Health Organization (WHO), countries have taken an aggressive measure to control the epidemic. About one-third of the world population is currently under lockdown to arrest the spread of this highly infectious disease.

Table 2 shows the list of a few geographical regions with their lockdown status. In the US, social distancing measures are enforced differently in different states and hence are reported separately for some key states most affected. The inclusion of Ohio is to show the disparity in the number of cases compared to a few other states which are most affected. Similarly, India is included to compare the number of cases when measures for social distancing were imposed.

**Table 2:** Social distancing measures by different countries (On April 9)

| Country | State/<br>Province | Lockdown |  | Single<br>day Peak<sup>2</sup> | Remarks |
| --- | --- | --- | --- | --- | --- |
|  |  | Date | Cases |  |  |
| US | New York | 22-Mar | 15168 | 7401 | New York State on PAUSE |
| US | New Jersey | 21-Mar | 1327 | 2474 | Stay at Home |
| US | Washington | 23-Mar | 2101 | 623 | Stay at Home |
| US | California | 19-Mar | 952 | 877 | Stay at Home |
| US | Ohio | 23-Mar | 444 |  | Stay at Home |
| China | Hubei | 2-Feb | 11177 | 6200 | Police enforced |
| Italy |  | 9-Mar | 9172 | 6557 | Police Enforced, Restricted domestic travel |
| Spain |  | 14-Mar | 6391 | 8271 | Police Enforced |
| France |  | 17-Mar | 7715 | 23060 | Strict nationwide lockdown, walk & travel restricted |
| Germany |  | 22-Mar | 24873 | 6933 | strict social distancing measures |
| UK |  | 23-Mar | 43847 | 5903 | Police Enforced, domestic/local travel allowed |
| India |  | 24-Mar | 536 | 809 | Police Enforced, *Janta* Curfew on 03/22 |

The effects of social distancing can also be pictorially seen in Fig. 3. In this figure, we can appreciate the shift in the number of infected cases in these key countries depending on the date of the lockdown. Thus, day 0 (*t*_0_) shows the cases when lockdown/stay-at-home was imposed. The orange shaded region, between *t*_*−*1_ and *t*_0_ shows the initial exponential growth typical of an epidemic, which continues through 14 days after the lockdown date to *t*_1_ = *t*_0_ + 14. The grey shaded region indicates the days from *t*_1_, when the effects of social distancing are expected to appear.

**Figure 3:**
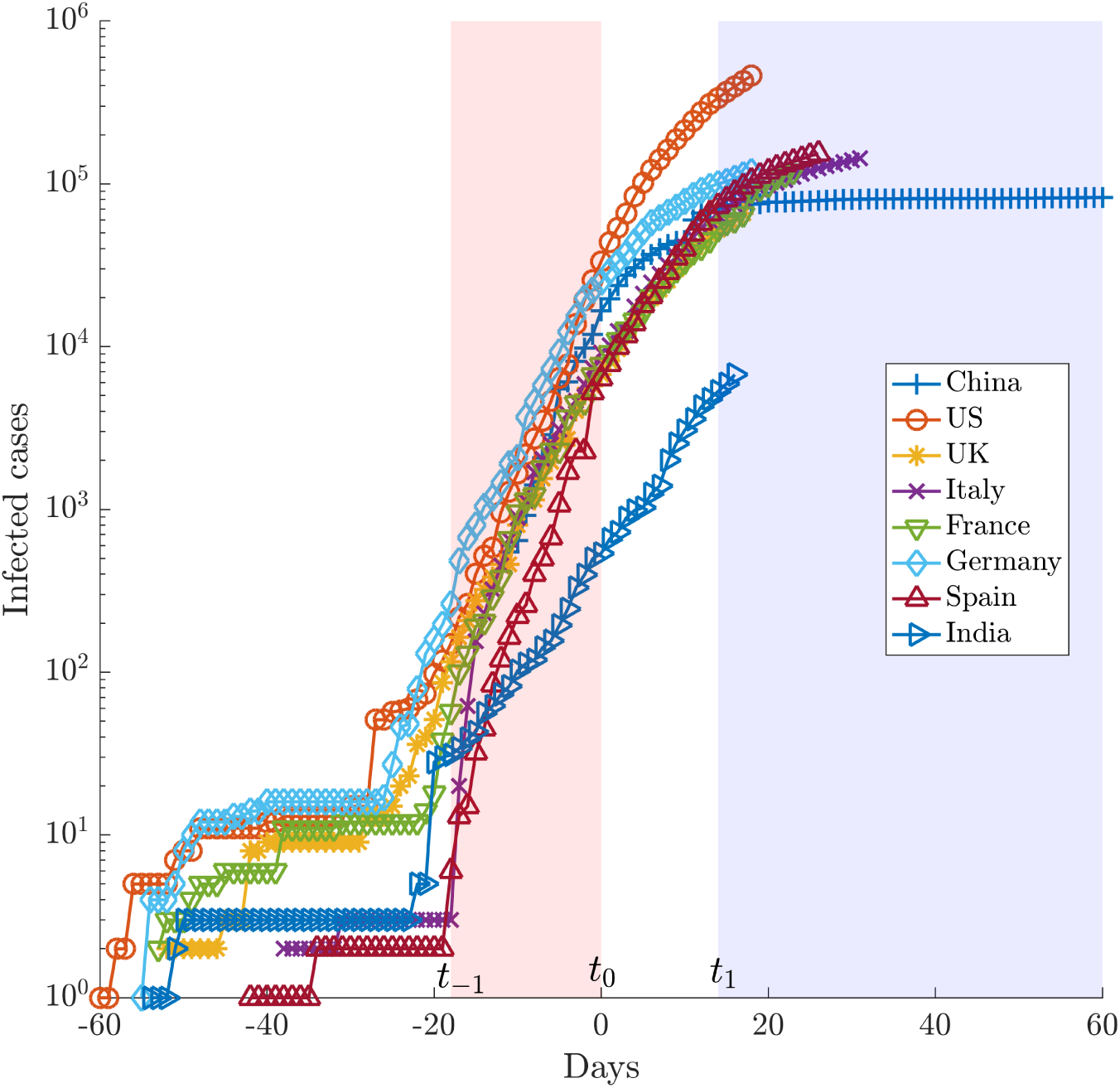
Effects of social distancing on flattening of the curve.*t*_*−*1_, *t*_0_ and *t*_1_ indicate beginning of the exponential growth, day on social distancing was enforced, and expected day to observe the effects of social distancing.

Particularly, we focus on the lockdown in China, where the curve has already flattened. The lockdown in Hubei province, the first epicenter of COVID-19, was enforced on Feb 2, when it had 11,177 cases. The cases in the Hubei province continued to rise till Feb 14 to reach a total of 54,406, with the total number of new cases reported on that day being about 6,200. Since then, there was a gradual drop in reported numbers of cases, and on Feb 15 and Feb 21, the new cases reported were 1,843 and 220, respectively. From Feb 2 to Feb 15, it took around 14 days for the lockdown to show its effect. The flattening of the curve took further 15 days. i.e. from Feb 15 to March 2. further discussions on this can be found in Ranjan[3].

It is known that the effects of social distancing become visible only after a few days from the lockdown. This is because the symptoms of the COVID-19 typically take some time to appear after getting infected from the Coronavirus SARS-CoV-2. The average incubation period is normally 5-6 days but it may take 2-14 days according to the Center for Disease Control and Prevention (CDC).

Most of the countries in Fig. 3 show flattening of the curve after *t*_1_, except for France, the UK and India. For France, recent spikes with single day spike of about 23000 has made an unusual change in the curve. For the UK and India also, there are no confirmed trends of decline in the acceleration rate. Further, it is important to note that for China, the flattening is relatively very sudden post lockdown while the deceleration in European countries is very slow. This aspect will be discussed further for introducing corrections to the predictions from models.

Modeling of an epidemic during its progress is very challenging as the parameters such as transmission rate, basic reproduction number etc. are not known. Values of these parameters are different for different geographical regions and depend on many social and environmental factors. Thus, understanding from the data from China, where the epidemiology is more established, cannot be directly translated to predict the epidemic size in other countries. Despite this, there have been several efforts to model the epidemic for different countries as an early estimate of the pandemic size can be very useful to design key strategies to tackle it.

The early stage of an epidemic, such as shown by the orange shaded region in Fig. 3, is relatively easy to model as the spread of disease follow an exponential curve. The modeling of later stages to predict the decline and eventual flattening of the curve is very challenging as more known parameters need to be included in the model. The inclusion of effects due to isolation and quarantine adds to the complication.

Typically, epidemiological models such as a logistic or SIR model is preferred for modeling the later stages. However, these models are highly dependent on initial conditions and underlying parameters, and an incorrect estimation of these can give completely different results. Further, modeling these only on data from an exponential curve can be misleading in estimating the final epidemic size. However, when there are signs of decline in the exponential growth (*t*_1_ in Fig. 3) and sufficiently long data is available, these models can be reliable for predictions within uncertainty limits. Though, waiting for the curves to show the sign of flattening, can be relatively late for making policy decisions. Nonetheless, a reliable estimate of the final epidemic size based on these data can be very helpful for taking necessary measures in terms of building adequate healthcare infrastructure and early preparation for the future. Further, it is known that despite the sign of decline, the epidemic curve can have a very long tail that may last up to a couple of months. Hence, despite a relatively small number of cases during the beginning of deceleration (at *t* = *t*_1_), the final epidemic size can be very large. These studies also provide insights about the timeline of a disease-free equilibrium.

In this work, we estimate the final epidemic size for some countries which has been hit badly due to this outbreak. Since, the results from epidemiological models can be dependent on its implementation to estimate parameters, initial condition, etc. even for same underlying data, we use three different models to compare the outcome. Apart from the logistic and simple SIR model, a generalized SEIR model is used that takes into account of quarantined and deceased patients. Further, we introduce a correction to the estimates from these models in order to account from slow decay of infections as seen in Fig. 3 for the US and other European countries.

Section 2.1 presents a brief discussion of these models: details of their implementation can be found in the corresponding references given. The corrections introduced to account for slow decay of infections compared to those observed in China are discussed in section 2.4. Section 3 shows the predictions for some key countries and a brief discussion is included. It is not possible to include the estimates for all the countries in this paper. However, it is expected that the reader can make use of these models, codes for which are available in the open-source, to estimate the final epidemic size for regions not included in this paper. As a caution, it should be mentioned that if the reported number of cases begins to exceed the predicted end-state systematically, then the epidemic will enter a new stage, and a new estimate needs to be made incorporating the effects of the changed parameters.

## 2 Materials and Methods

### 2.1 Epidemiological Models

#### 2.1.1 Exponential Model

It is known that during early stages of a pandemic, the growth in the number of diagnosed infections with time is exponential. Thus, if the number of diagnosed infections *C*(*t*) over time *t* is known, one can find the growth rate *r*,

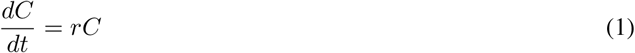

Integrating, we get-

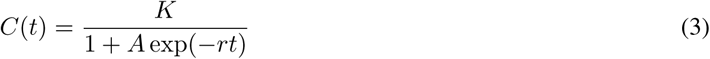

Where *C*_0_ is constant that can be obtained by fitting the curve with the available data. In this model, *C* is taken as the cumulative number of diagnoses (including individuals who have recovered(*R*) or deceased(*D*). This model can successfully predict the early stages of the epidemic such as between *t* _*−*1_ and *t*_0_ (and sometimes up to *t*_1_) in Fig. 3. The results from this model are not included in this study. Ranjan [3] has discussed this model for prediction of early stages in COVID-19 epidemic in India.

#### 2.1.2 Logistic Model

While the exponential model predicts the initial growth of a pandemic, it does not account for eventual decay and flattening out of the curve. Logistic model, on the other hand, may predict the eventual decay but may fail in initial stages. In Logistic model, the growth is given by[4]:

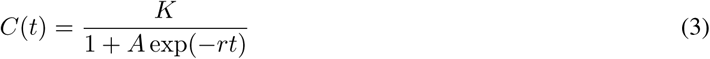

where *A* = (*K/C*_0_) *−* 1, assuming *K »C*_0_ and hence *A»*1. For small time *t*, this approximates the exponential model. In the present study, non-linear fit for the regression model has been obtained using the Least Squares approach.

#### 2.1.3 SIR Model

Susceptible-Infectious-Recovered (SIR) model is a compartmental model that accounts for number of susceptibles *S*, number of infectious *I*, and the number of recovered or deceased (or immune) individuals *R*. Their distributions can be given as following[5] :

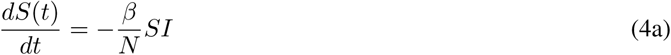

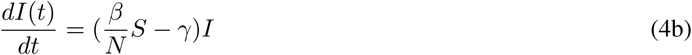

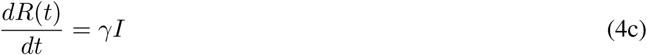

where *β* is the transmission rate, and *γ* is the average recovery rate. Note that here constant *N* is not the population of the country but population composed of susceptibles (*S*), infected (*I*) and recovered (*R*). In the present model, at any time the total population *N* = *S* + *I* + *R* remains constant as 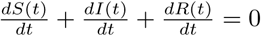 Initially, *N* is approximately equal to *S* as *I* is very small. In an outbreak, typically *N* will increase every day because more people may get affected due to local outbreaks. However, with quarantine and isolation, this number will slowly become constant. Hence, this SIR model, where *N* is assumed constant, is valid provided measures are taken to ensure *N* does not increase much with time. In the present situation, most of the countries have taken extensive measures to impose rigorous social distancing and thus increase in *N* may not be significant. This model assumes equally likely recovery of everyone affected and hence does not consider factors like age, morbidity etc.

We assume a disease-free equilibrium (DFE) for a completely susceptible population i.e. the final number of infected people is zero. In the current approach, the initial guess of *γ* and *β* are obtained by setting *R*(0) = 0 and then the SIR equations are solved. Details of the implementation can be found in Batista[6]. For this purpose, the *fitViruscv19v3* code developed by Batista[7] is used. In this, to estimate the parameters and initial values, the differences between the actual and predicted number of cases with time are minimized using MATLAB function *fminsearch*. The SIR equations are integrated using MATLAB function *ode45*.

#### 2.1.4 Generalized SEIR (SEIQRDP) Model

The generalized SEIR Model as proposed in Reference[8] includes many more states than SIR model, namely susceptible cases (*S*), insusceptible cases (*P*), exposed cases (*E*), infectious cases (*I*), quarantined cases (*Q*; confirmed and infected), recovered cases (*R*) and deceased cases (*D*). The governing equations for each of them are given in 2.1.4. The coefficients *α, β, γ*^*−*1^, *δ*^*−*1^, *λ*(*t*), *κ*(*t*) represent the protection rate, infection rate, average latent time, average quarantine time, cure rate, and mortality rate, separately. Among these the cure rate *λ*(*t*) = *λ*_0_(1*−* exp(*− λ*_1_*t*)) and mortality rate *κ*(*t*) = *κ*_0_ exp(*−κ*_1_*t*) are time dependent, with the former increasing with time while the latter first increases and then rapidly decreases. *λ*_0_, *λ*_1_, *κ*_0_, *κ*_1_ are constant parameters.

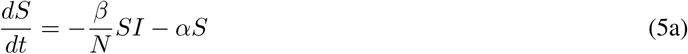

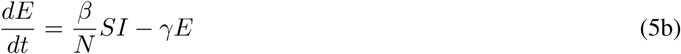

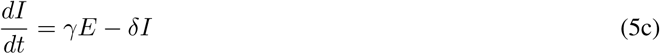

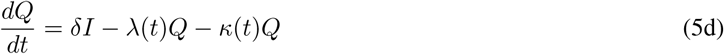

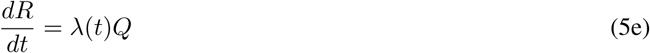

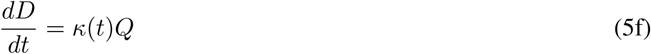

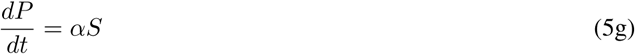

Note that, because of the inclusion of insusceptible cases, here *N* = *S* + *P* + *E* + *I* + *Q* + *R* + *D* represent the total population of a given geographical region unlike the SIR model. The details of this implementation are given in [8]. Open source Matlab code developed by [9] is used for this purpose. Briefly, time histories of the number of quarantined *Q*(*t*), recovered *R*(*t*), and deceased *D*(*t*) cases are used to estimate the parameters using least-squared nonlinear curve-fitting tool *lsqcurvefit*. Equations are solved using 4^*th*^ order Runge-Kutta method.

### 2.2 Data

The data for the models have been taken from ‘Johns Hopkins University Coronavirus Data Stream’ that combines World Health Organization (WHO) and CDC case data. Specifically, the time-series data from https://github.com/CSSEGISandData/COVID-19 till April 11, 2020 has been used for current modeling purposes. These data are separately available for all the global regions, as well as for states and counties in the US from date January 22, 2020. The time-series global data include total confirmed (*C*), recovered (*R*) and deceased (*D*) cases for all the affected countries. It is assumed that all the confirmed cases are sent to quarantine. Therefore, the number of quarantined *Q*(*t*) cases at any time are given by *C*(*t*) *− R*(*t*) *− D*(*t*). For the US, time histories of only confirmed (*C*) and deceased (*D*) cases are available for all the states and counties. In these cases, the fit in the SEIQRDP model is obtained using the data of quarantined plus recovered cases (*Q*(*t*) + *R*(*t*) = *C*(*t*) *− D*(*t*)).

### 2.3 Validity of the Models

We first show validity and limitation of these models by taking the data for Hubei and Hunan in China. Predictions using all these models are made using part of the available data and then are compared with actual data. Figure 4 shows these results. It may be noticed that if the data included for modeling come only from the acceleration region (exponential growth part) of the curve, the predictions of eventual epidemic size are very different from the actual numbers. Further, predictions by different models differ from one another in such cases. This highlights the convergence issue in estimation of modeling parameters. The predictions improve when part of the flattened curve is also included for the estimation of parameters in the model. The predictions by models SIR and SEIQRDP are very close to each other. The logistic model consistently underpredicts the final epidemic size.

**Figure 4:**
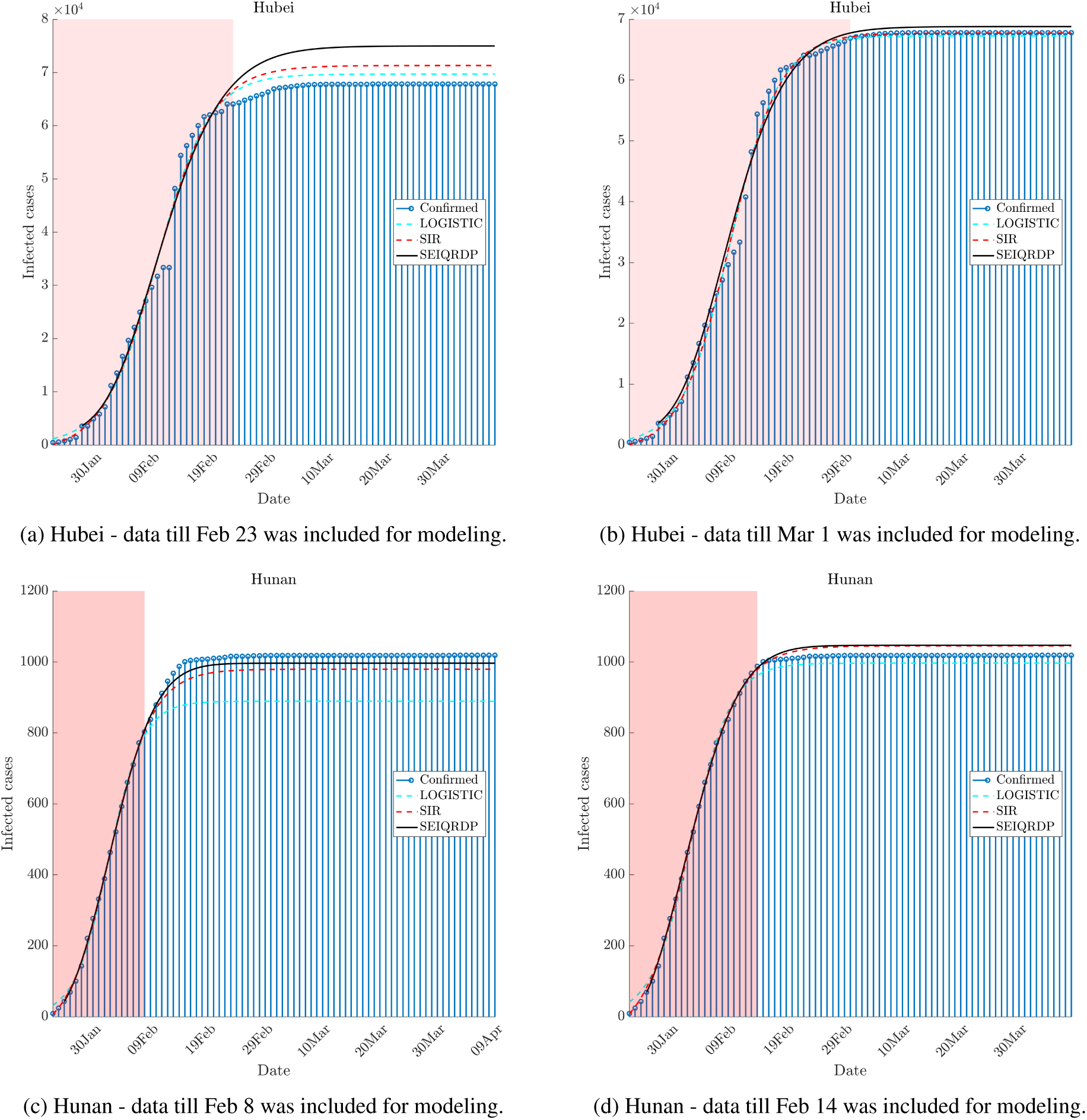
Predictions vs actual using epidemiological models. Shaded region show the data that has been used for predictions. Accuracy increases when the data for the initial flat curve is also included for predicting model parameters.

### 2.4 Corrections due to slow decay of infections

The infection rate predicted by all the above models follow almost a normal distribution with 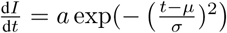, with *μ* represents the day of peak cases, *σ* is spread around this day from the beginning of the epidemic to the end, and *a* is the constant. Because of the availability of limited data, predicting both - day of the peak (mean) as well as the spread (variance) is challenging for every model. Further, even if the day of the peak is known the decline rate is often not correctly predicted giving an incorrect estimation of the final epidemic size. Because of the symmetric distribution of the curve, the decline rate is predicted as negative of the acceleration. This is often not true as observed from the COVID-19 data of Italy and Spain, where the decline is very slow. In order to illustrate this point, we show the total infection as well as the rate of infection for Italy in Fig. 5. The predictions by the three models are also shown. All the three fitted curves show underprediction of total confirmed cases from April 8 despite using data till April 11 for modeling. If we look at the infection rate, we note that the decline rate given by Gaussian-like distribution is much higher than actual decline leading to a lower estimate of the epidemic size. In order to account for this difference, we introduce a new parameter *η*, which changes the variance of this distribution for predictions after the peak. Briefly, we fit a normal distribution after getting predictions from SEIQRDP model, and then adjust the spread to make the decline rate realistic. Hence, the new distribution is given by 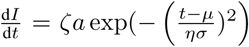, where the factor *ζ* = *a*_1_*/a* ensures that number of infections on the peak day remains unchanged. The dash-dot magenta curve shows this new distribution which is much closer to the actual values. The value of *η* is obtained by fitting the curve to Italy and used for subsequent predictions. This corrected model is designated as SEIQRDP(C). Predictions from this model give the upper limit of the predicted cases. Note that, this adjustment was not needed for fitting data of China as the flattening of curve in China was very fast compared to what is seen in the recent past for European countries (see Fig. 3).

**Figure 5:**
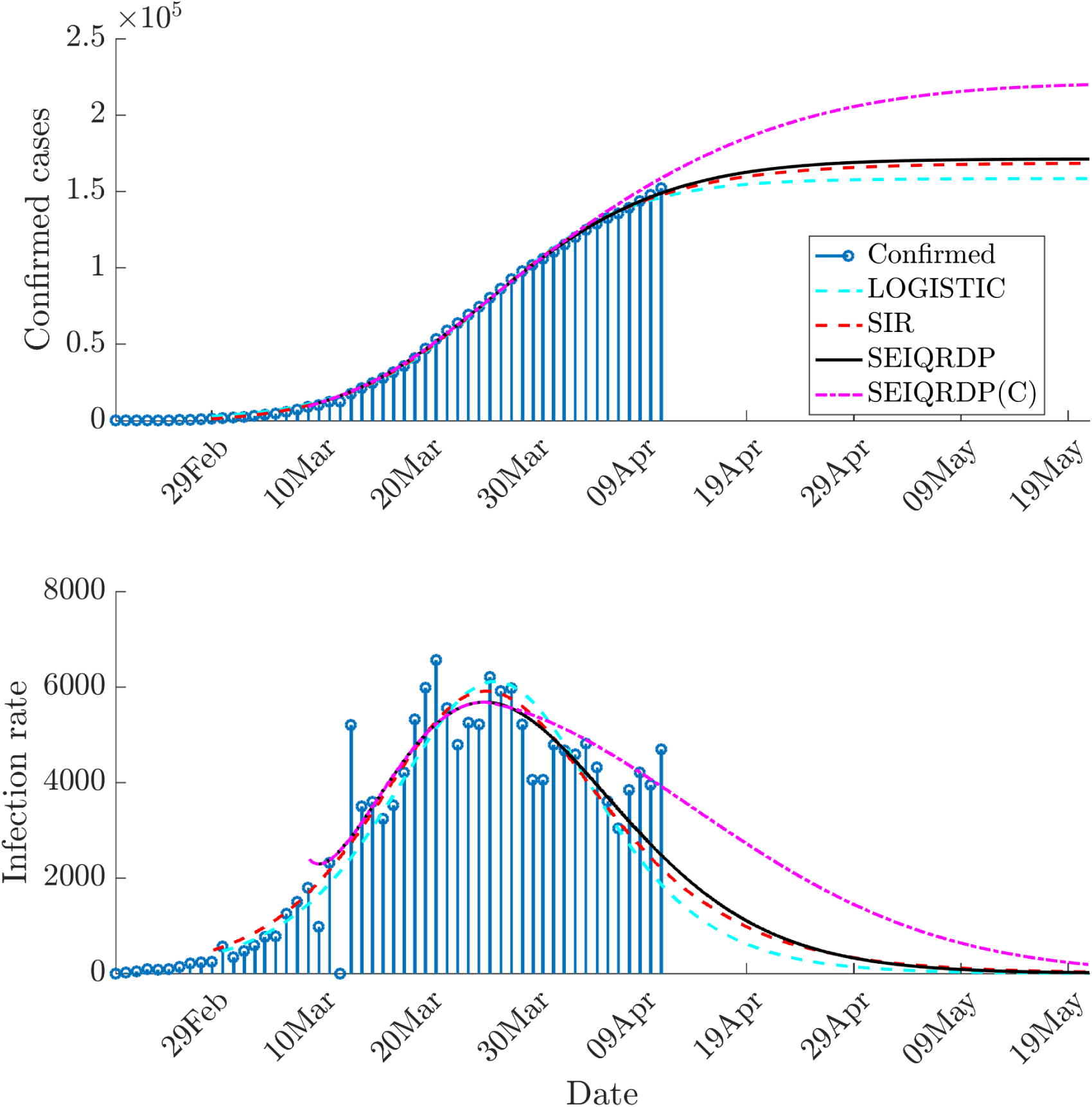
Prediction for Italy. Note the decline in the infection is much faster in the predicted curves from logistic, SIR, SEIQRDP models compared to actual cases. Corrected predictions with SEIQRDP(C) brings the predictions closer to actual values and gives the upper limit of estimated total cases.

## 3 Predictions

Now we show the predictions of the final epidemic size for regions most affected. Though not shown for individual cases, it is ensured in every case that the fits are statistically significant with *R*^2^ *>* 0.97 and p-value very close to zero.

Predictions for the US are shown in Fig. 6. Both SIR and SEIQRDP models both give very close predictions of the final epidemic size of around 0.85 million, indicating good convergence of model parameters. As expected, the predicted size from the logistic model is much smaller than this value. Further, flattening of the curve begins in the third week of April. The corrections due to adjustment of deceleration gives the final estimate about 1.09 million. The final epidemic size for all the regions discussed in this paper is given in Table 3. Figure 6(b) also shows predictions for deaths using SEIQRDP. It is estimated that total deaths by the end of the epidemic could be around 33000, which is around 4% of total estimated cases. This is a very optimistic case as countries like Italy and France, have respectively reported 13% and 11% deaths per total infections.

**Table 3:** Final epidemic size

| Country | State | Model |  |  | Remarks |
| --- | --- | --- | --- | --- | --- |
|  |  | SIR | SEIQRDP | SEIQRDP(C) |  |
| US |  | 846401 | 853720 | 1093106 |  |
| US | New York | 319842 | 314553 | 403404 |  |
| US | New Jersey | 72531 | 74077 | 94804 |  |
| US | Michigan | 30494 | 31175 | 40365 |  |
| US | California | 31118 | 32701 | 42365 |  |
| US | Washington | 13424 | 13906 | 18331 |  |
| US | Ohio | 8658 | 8533 | 10945 |  |
| Italy |  | 168613 | 171239 | 221311 |  |
| Spain |  | 187097 | 185822 | 241136 |  |
| Germany |  | 145968 | 147535 | 189767 |  |
| UK |  | 131104 | 206446 | 264533 | Not converged |
| France |  | 155527 | 395311 | 493224 | Not reliable |
| Iran |  | 100970 | 141009 | 182558 | Not converged |
| India |  | 16916 | 25246 | 36323 | Not converged |

**Figure 6:**
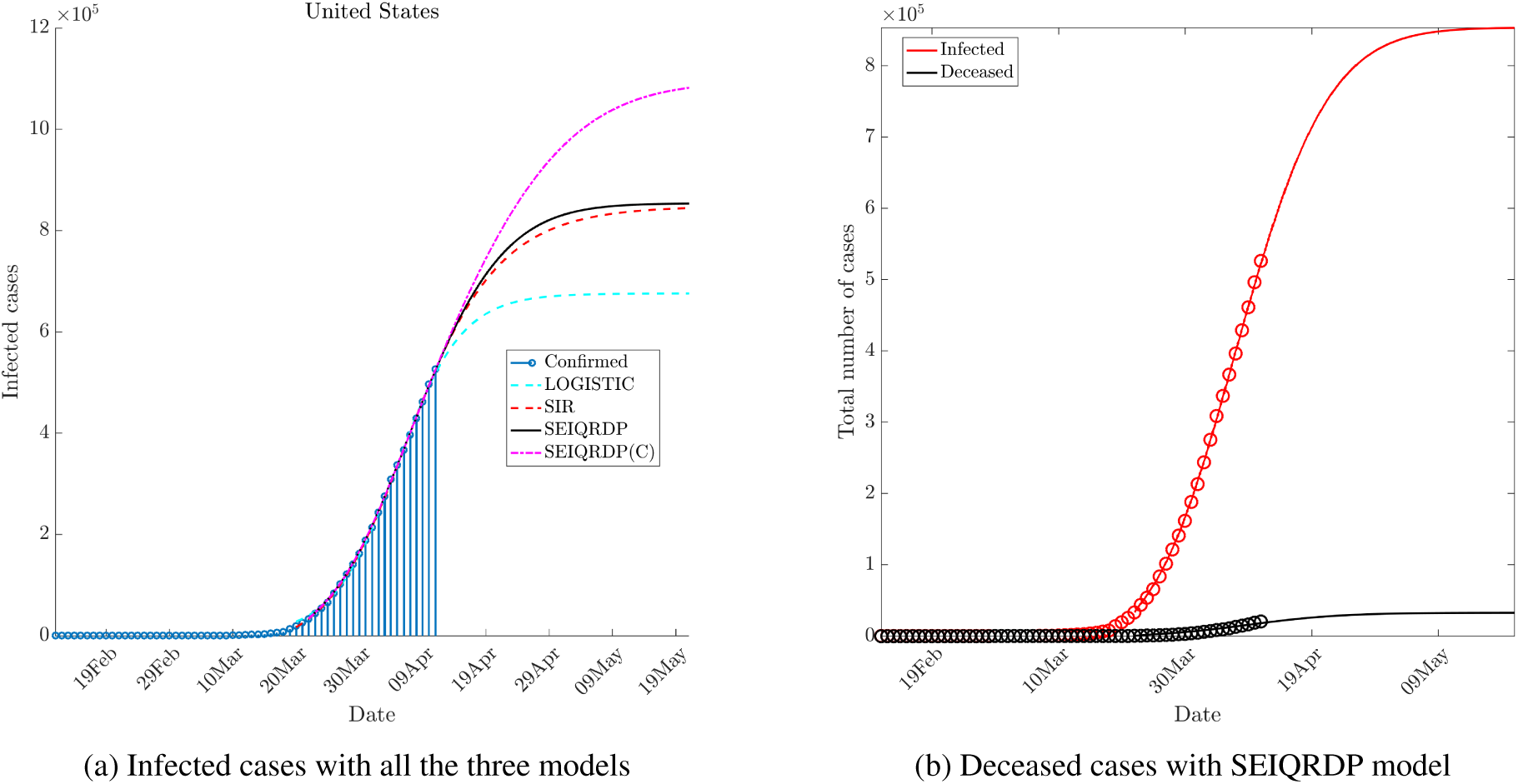
Prediction for the US.

Fig. 7 shows the infection rate with the SIR model. Though the estimated decline begins on April 6, the curve shows a very long tail (even without correction) till the middle of May for zero new infection. As shown in Table 2, there is a great disparity in the number of cases in different states in the US. New York alone accounts for 30% of the total cases in the US. Thus, we show predictions for some key states in the US in Fig. 8. It is estimated that the number of cases in New York could rise to around 0.4 million at the end of epidemic. Estimations for New Jersey, California, Washington, Michigan, and Ohio are also shown and are also tabulated in Table 3. For some of the states, such as New Jersey, California and Michigan, SIR and SEIQRDP models give slightly different estimates. However, as shown in table 2, these estimates are not very far from each other, and give a range of expected final size. The total number of cases in Ohio may limit to 10945 according to predictions. This may be due to early action of social distancing measures compare to a few other states (see Table 2. Of course, these predictions depend on the compliance level of the social distancing measures so that governing parameters in the model do not change significantly.

**Figure 7:**
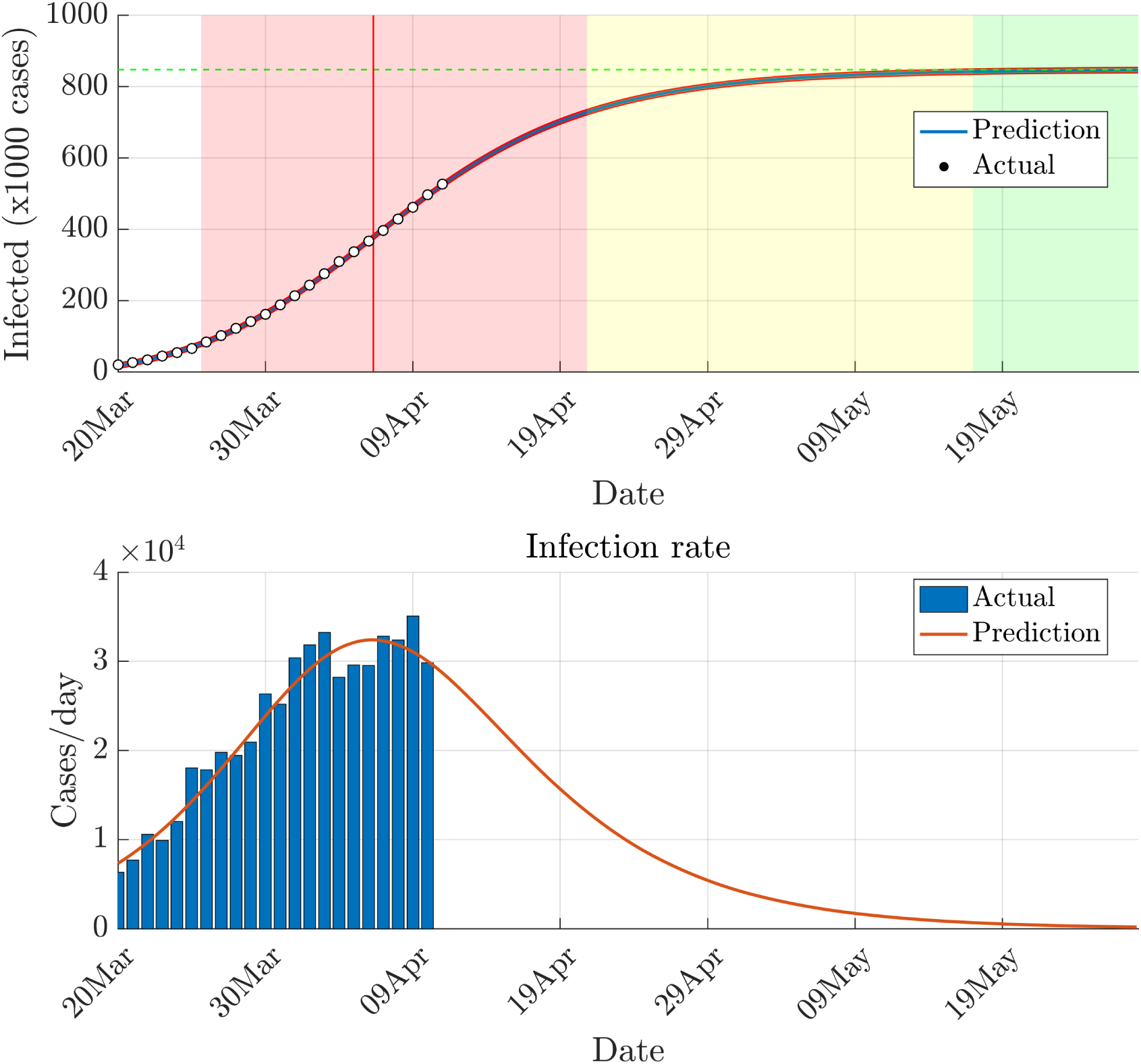
Prediction with the SIR model for the US. Top panel shows different epidemic phases based on this model. White, red, yellow and green regions indicate initial exponential growth, fast growth (with positive and negative phase separated by red vertical line), asymptotic slow growth and curve flattening, respectively.

**Figure 8:**
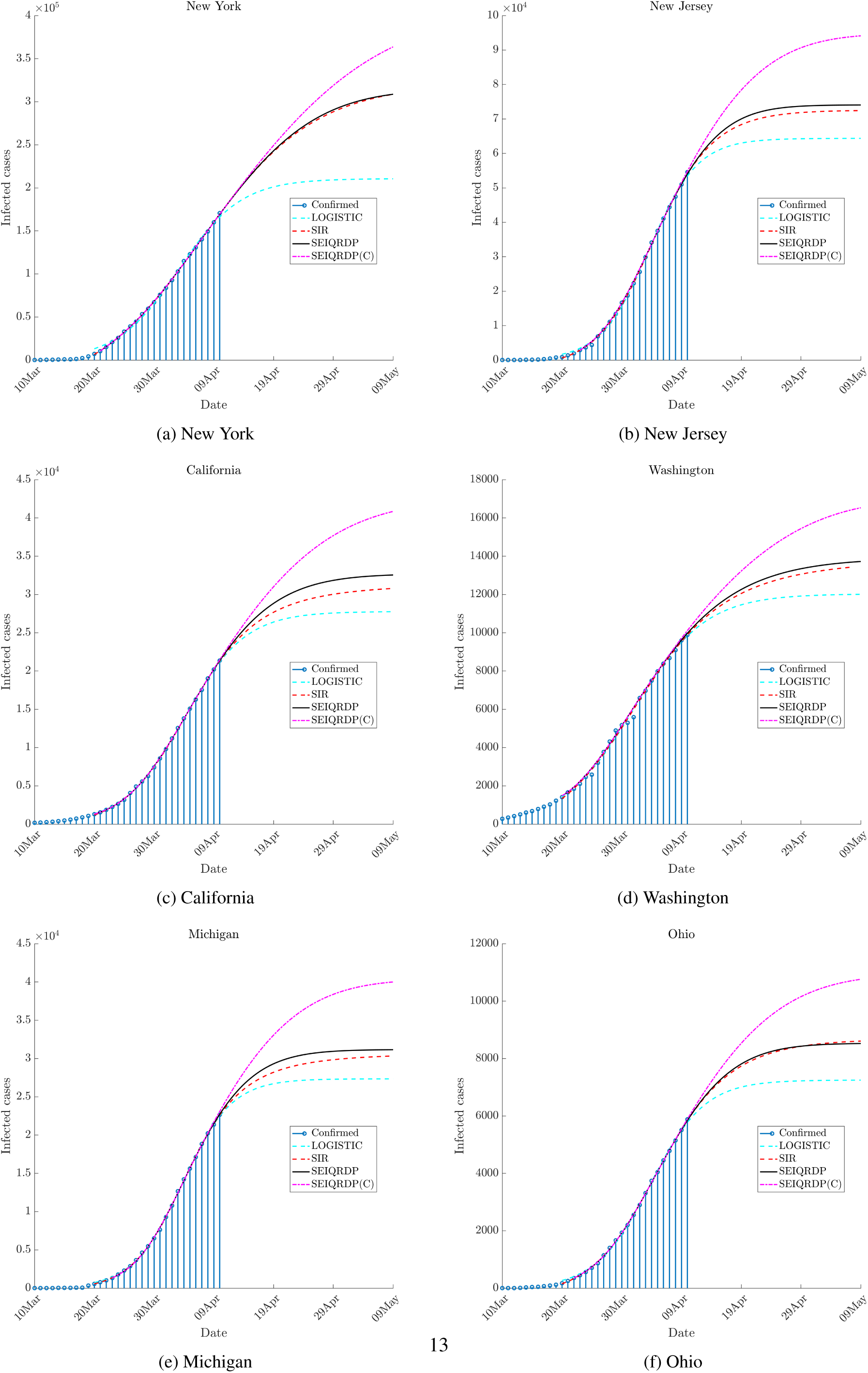
Prediction with all the models for some key states in the US.

The estimate from the US as a whole should be taken with following considerations. Unlike countries like India and several other European countries, different states in the US have different norms for enforcing social distancing. In fact, many states where the COVID-19 outbreak is not yet severe, have not strict actions to enforce social distancing measures. Further, domestic travel is still allowed in the US unlike India, Italy and France. Therefore, it is possible that while key states like New York and New Jersey show decline in the infection rate due to social distancing, other states where there is a small number of cases as of now may start showing fast exponential growth due to lack of sufficient measures. Therefore, the numbers reported in Table 3 for the US could still be optimistic.

Lastly, we show the predictions for countries worst affected by the epidemic in Fig. 9. Good convergence is obtained for Spain, Italy, and Germany with an estimated final epidemic size of 0.23, 0.21 and 0.184 million respectively. On the other hand, the estimates for France, UK, Iran and India show convergence issues with a disparity in predictions with SIR and SEIQRDP models. This behavior is expected as discussed earlier as the curves in these countries are still in accelerating stages due to recent outbreaks (shown earlier in Fig. 3). Among these, the predictions for France are not at all trustworthy as the recent big jump in the number of cases makes the convergence of model parameter very difficult. Results with the UK, Iran and India are also not converged but gives an order-of-magnitude estimation of final epidemic size. A very low estimate for India may be due to early measures of international flight suspension and strict lockdown before stage-3. So far, only a few small cluster community transmissions are reported in India.

**Figure 9:**
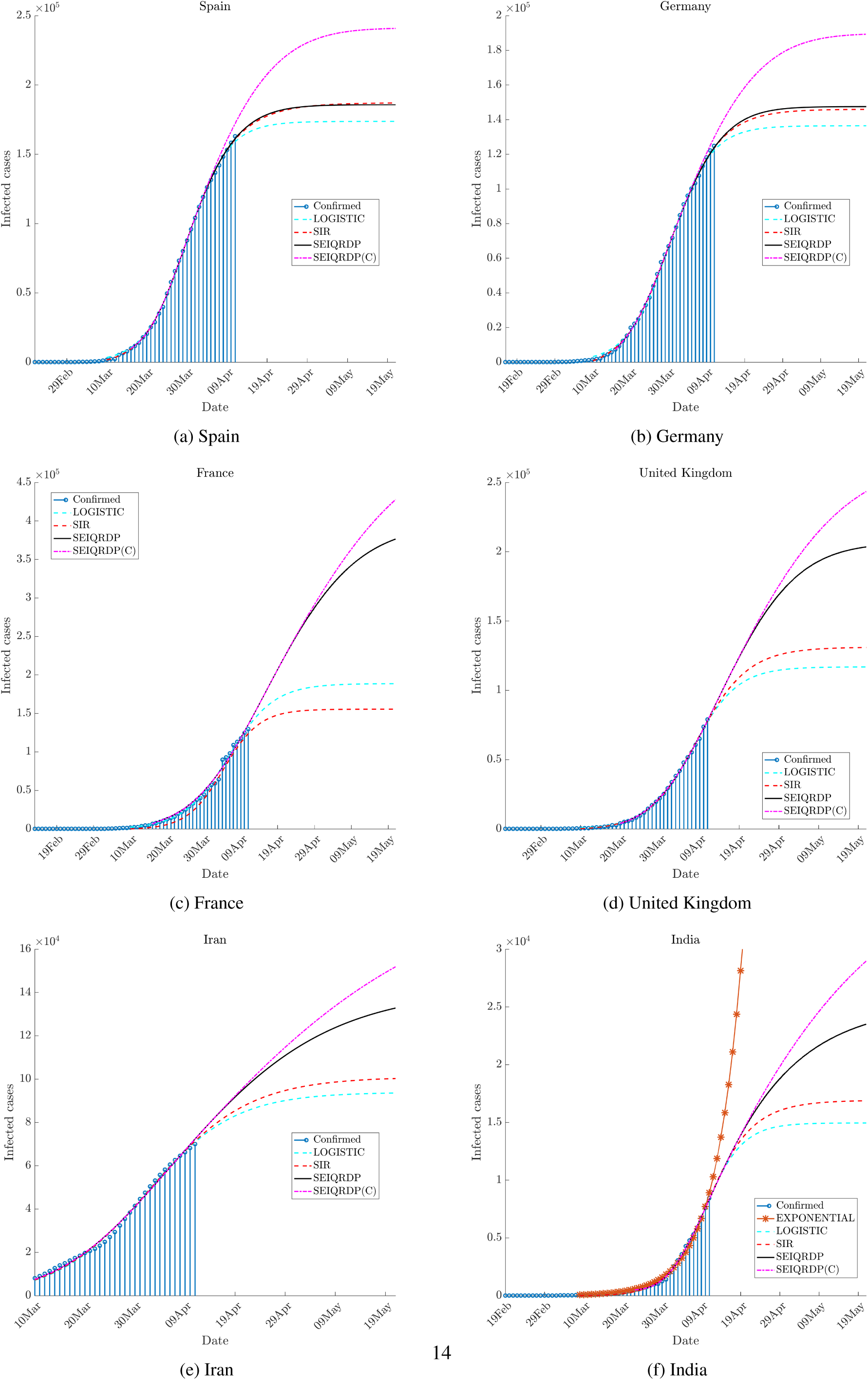
Prediction for countries most affected by Epidemic, and India.

The final epidemic size of the entire world is difficult to estimate without getting individual estimates of all the countries. This is because the total number of cases per day still show an exponential growth trend worldwide as shown in Fig. 2(a).

## 4 Conclusions

Three epidemiological models-logistic, SIR and generalized SEIR models are used to make predictions for the final epidemic size of COVID-19. Both SIR and generalized SEIR models give similar predictions for regions with signs of flattening of COVID-19 curve, while the logistic model consistently gives lower estimates. Similar estimates with SIR and generalized SEIR models, which use very different parameters and initial values, indicate that the underlying data are reliable to give convergence of model parameters. Thus, the estimations with these models could be more reliable than early estimates in the literature that has used data only from the acceleration phase. However, symmetric distributions of the infection rate predicted by the models around the peak value are not realistic as the decay rate in infections is generally much slower than acceleration as seen from the recent data from Italy and other European countries. Hence, a correction based on the change in the variance of the normal distribution is introduced to account for this factor. This generally provides the upper limit of these estimates. Finally, these models are used to estimate the final epidemic size for key countries where the epidemic outbreak is severe. The epidemic size for a few states in the US are also reported. A good convergence for France, UK, Iran, and India could not be obtained due to recent outbreaks. However, it is hoped that these models give a reasonable estimate for range of final total infections and can be used with confidence for regions where decline or at least no increase in the infection rate is visible.

## Data Availability

The data that support the findings of this study are publicly available.
https://github.com/CSSEGISandData/COVID-19

## 5 Acknowledgment

Author would like to acknowledge Dr. Deepti Chugh (The Ohio State University) and Dr. Sudheendra N R Rao (Scientific Advisor, Organization for Rare Diseases India) for their useful inputs in this work.

## Footnotes

2 Data till April 9. Hubei has anomaly in data as no new case was reported on Feb 12 and abruptly high number of cases (∼ 14840) was reported on the following day.

